# From SARS-CoV-2 infection to COVID-19 disease: a proposed mechanism for viral spread to the lower airway based on *in silico* estimation of virion flow rates

**DOI:** 10.1101/2020.12.19.20248544

**Authors:** Saikat Basu, Arijit Chakravarty

## Abstract

While the nasopharynx in the upper respiratory airway is the dominant initial infection site for SARS-CoV-2, the physiologic mechanism that launches the infection in the lower airway is still not well-understood. Based on the rapidity with which SARS-CoV-2 infection progresses to the lungs, it has been conjectured that the nasopharynx acts as the seeding zone for subsequent contamination of the lower airway via aspiration of virus-laden boluses of nasopharyngeal fluids. In this study, we examine the plausibility of this proposed mechanism. To this end, we have developed computational fluid mechanics models of the inhalation process in two medical imaging based airway reconstructions and have quantified the nasopharyngeal liquid volume ingested into the lower airspace during each aspiration. The numerical predictions are validated by comparing the number of projected aspirations (approximately 2 – 4) during an eight-hour sleep cycle with prior observational findings of 3 aspirations in human subjects. Extending the numerical trends on aspiration volume to earlier records on aspiration frequency for the entire day indicates a total aspirated nasopharyngeal liquid volume of 0.3 – 0.76 ml per day. We then used sputum assessment data from hospitalized COVID-19 patients to estimate the number of virions that are transmitted daily to the lungs via nasopharyngeal liquid boluses. For mean sputum viral load, our modeling projects that the number of virions penetrating to the lower airway per day will range over 2.1 × 10^6^ – 5.3 × 10^6^; for peak viral load, the corresponding number of penetrating virions hovers between 7.1 × 10^8^ – 17.9 × 10^8^. These findings fill in a key piece of the mechanistic puzzle of the progression from SARS-CoV-2 infection of the nasopharynx to the development of COVID-19 disease within a patient, and point to dysphagia as a potential underlying risk factor for COVID-19. The findings also have significant practical implications in the design of COVID-19 prophylactics and therapeutics that aim to constrain the pathogenic progress of the disease within the limits of the upper airway.

## 1 Introduction

Severe acute respiratory syndrome coronavirus 2 (SARS-CoV-2), the causative agent for coronavirus disease 2019 (COVID-19), has been linked^1,2^ to a remarkable pattern of relatively high infectivity in ciliated epithelial cells along the nasal passage lining in the upper airway, moderate infectivity in cells lining the throat and bronchia, and relatively low infectivity in lung cells. Such viral tropism is governed by the abundance of angiotensin-converting enzyme 2 (ACE2), a single-pass type I membrane protein that is exploited by the viral spike protein binding as a gateway for cellular entry. ACE2 is abundant on ciliated epithelial cells, but is highly expressed in only a smaller subset of the alveolar cells in the lower airway. The findings are nonetheless for *in vitro* samples; virus-laden droplets deposited along the anterior nasal airway might not be so effective at launching an infection despite the presence of ciliated cells, as the thicker mucus layer there provides some level of protection against virus invasion and infection^3^. Hence, the *nasopharynx*, which is the region in the upper airway posterior to the septum comprising the superior portion of the pharynx, has been postulated to be the main initial infection site for SARS-CoV-2. Efficacy of nasopharyngeal swabs over oropharyngeal swabs for accurate detection^4^ of COVID-positive cases supports this hypothesis. Based on the brisk pace at which lower airway infections ensue following the emergence of initial symptoms^1^, it may be conjectured that the nasopharynx (marked in Fig. 1, see Panels (a)-(f)) acts as the seeding zone for subsequent spread of the disease to the lungs via lower airway aspiration of virus-laden boluses of nasopharyngeal fluids.

**Figure 1.**
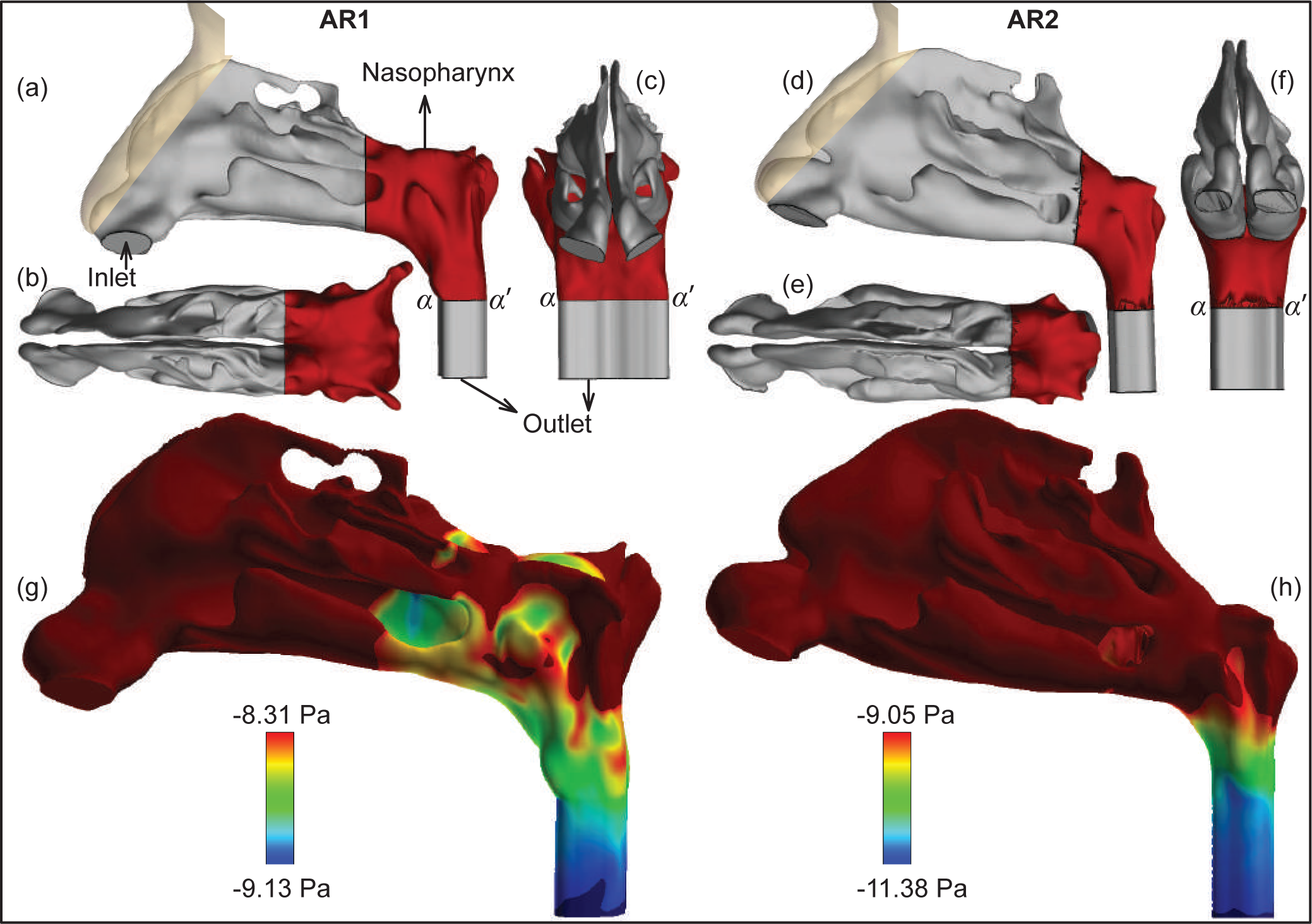
Panels (a), (b), and (c) respectively show the sagittal, axial, and coronal views of the CT-based upper airway domains for anatomic reconstruction 1 (AR1); Panels (d), (e), (f) show the corresponding views for anatomic reconstruction 2 (AR2). Nasopharynx is marked in red. Additionally, *α* − *α* ^*′*^ shows the location of the wetted perimeter used while estimating the hydraulic radius (see Section 2.2.3). Panel (g) depicts the wall pressure map in AR1, for simulated inhalation of 15 L/min. Panel (g) shows the wall pressure map for AR1, for simulated inhalation of 15 L/min. The pressure color-maps for (g) and (h) set the limiting contour colors at the *p*_*n*_ (averaged wall pressure at nasopharynx) and *p*_*o*_ (averaged pressure at outlet) values that are extracted from the simulated data, in each case. The data post-processing is done on FieldView, under software license provided through the University Partners Program.

While this conjectured mechanism is superficially plausible, a key unanswered question is whether the rate of virion flow from the initial site of infection to the lungs would be sufficiently high to account for a second infection site. In this report, we have combined earlier data on aspiration trends^5,6^ with virological assessments of sputum in COVID-positive patients^7^ and computational findings on the flow physics variables in anatomically realistic airway models^8^ – to quantify bolus-borne virion transmission rates from the nasopharynx into the lower airway.

## 2 Materials and Methods

### 2.1 Frequency and quantification of pharyngeal aspiration

Aspiration of upper airway secretions acts as a major carrier of pathogens to the lower airway, and the phenomenon, fortunately enough given the current pandemic, has been studied in great detail over the last few decades. As reported in the late-1990s^5^, aspirated pharyngeal liquid volume during sleep ranges from 0.011 ml to 0.129 ml, measured through tracking mildly-radioactive tracers after the subjects wake up. Further introspection of the data clearly shows that the maximum data-point in the reported range is a statistical outlier. Including the maximum-reported volume in the analysis, the mean aspirated volume comes out to be 0.0345 ml and the median is 0.0215 ml. Excluding the outlier, the mean volume revises to 0.021 ml and the median volume adjusts to 0.020 ml. The study was based on a total of 10 normal subjects.

Further, while evaluating swallowing mechanisms, it has been reported^6^ that for bolus volumes 5 ml or less, aspirations happen during 13% of swallows; and for bolus volumes 10 ml or less, aspirations happen during 11% of swallows. Boluses smaller than 10 ml are associated with silent aspiration^9^ and presumably are the major sources of pathogen-carriers to the deep lungs, and with averaging the reported data, 12% of the swallowing actions (for 10 ml or smaller bolus volumes) should result in aspiration.

Finally, earlier findings^10^ suggest that a typical person will swallow 500 – 700 times during a day and 24 times during sleep (assuming a standard eight-hour sleep cycle). These numbers thus indicate that a subject will aspirate approximately 12% of 500 – 700 times, i.e. 60 – 84 times during the day, and 12% of 24 times, i.e. approximately 3 times during sleep.

### 2.2 Development of anatomically realistic computational fluid mechanics models

Allometric relations^11^ show that the minute inhalation is approximately 14.5 – 20.0 L/min for a 65-kg male and 8.8 – 22.4 L/min for a 65-kg female, both for gentle steady breathing. For simplicity, in this study, we have simulated inhalation at only 15 L/min; the process can be modeled using viscous-laminar steady state flow physics schemes^12–21^.

#### 2.2.1 Anatomic airway reconstructions

The *in silico* anatomic geometries were reconstructed from medical-grade computed tomography (CT) scans sourced from existing de-identified imaging data from two CT-normal subjects. Use of the archived records was approved with exempt status by the Institutional Review Board of the University of North Carolina at Chapel Hill, with the informed consent requirement waived for retrospective computational use. The test subjects include a 61 year-old female (subject for anatomic reconstruction 1, or AR1) and a 37 year-old female (subject for anatomic reconstruction 2, or AR2). As for imaging resolution, the CT slices were collected at coronal depth increments of 0.4 mm. The anatomic airspace was extracted from the scans over a delineation range of -1024 to -300 Hounsfield units, the process was complemented by careful hand-editing of the selected pixels for anatomic accuracy. Subsequently, the reconstructed geometries were spatially meshed into minute volume elements. As per established mesh refinement protocols^22,23^, each computational grid in this study has more than 4 million unstructured, graded tetrahedral elements (namely, 4.54 million in AR1, 4.89 million in AR2).

#### 2.2.2 Simulating inhalation

The viscous-laminar flow simulations of the inhalation process were carried out using a segregated solver on ANSYS Fluent, with SIMPLEC pressure-velocity coupling and second-order upwind spatial discretization. We monitored the solution convergence by minimizing the mass continuity and velocity component residuals, and by stabilizing the mass flow rate and static pressure at the airflow outlets. For the pressure-driven flow solutions, typical run-time for 5000 iterations was 5 – 6 hours through 4-processor based parallel computations executed at 4.0 GHz speed. We assumed the air density to be 1.204 kg/m^3^ and the dynamic viscosity of air to be 1.825 × 10^−5^ Pa.s.

We enforced the following boundary conditions in the flow simulations: zero velocity (*no slip*) at the internal airway walls i.e. at the tissues and cartilages enclosing the airway; zero pressure at nostril openings, which acted as the pressure-inlet zones; and negative pressure at the airflow outlet at the base of the nasopharynx, which acted as the pressure-outlet zone. See Fig. 1 for the relative locations of the anatomic regions.

#### 2.2.3 Theoretical estimation of aspirated nasopharyngeal liquid volume

For an instantaneous pathogenic event such as pharyngeal aspiration, the transport of the nasopharyngeal liquids downstream to the lower airway can be mathematically described through a reduced-order model of steady unidirectional flow. With the assumption of axial symmetry in the airway conduit and *no slip* boundary condition at the walls, integrating the Navier-Stokes equation for momentum conservation results in

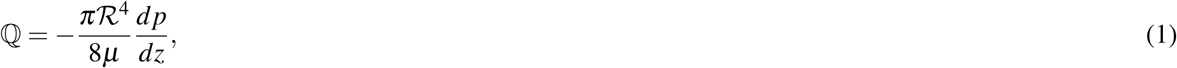

where ℚ is the instantaneous aspirated volume, ℛ is the hydraulic radius (cross-section of conduit divided by the perimeter at the nasopharyngeal base), *µ* is the sputum viscosity (quantified at 4.59 Poise for mucopurulent^24^), and *dp/dz* is the spatial rate of pressure gradient in the streamwise direction. To adapt the formulation to the present problem, we have post-processed the simulations to extract the averaged wall pressure at the nasopharynx (*p*_*n*_), since the bolus would typically arise from the shearing action along nasopharyngeal surfaces. We have also extracted the averaged pressure (*p*_*o*_) at the outlet (see Fig. 1), located 20 mm (streamwise Δ*Z*, so as to ensure full flow development in the simulations) below the nasopharyngeal base. With Δ*P* = *p*_*o*_ − *p*_*n*_, the gradient rate *dp/dz* can therefore be approximated to simply Δ*P/*Δ*Z*.

### 2.3 Data on viral loading

SARS-CoV-2 is a single-stranded RNA virus, and virological assessments^7^ performed on the sputum of hospitalized COVID-19 patients indicate a mean viral load of *𝒱*_ave_ = 7 × 10^6^ RNA copies/ml of oral fluid. The peak load was *𝒱*_peak_ = 2.35 × 10^9^ copies/ml.

## 3 Results

### 3.1 Computational prediction of shear-generated, pressure-driven nasopharyngeal bolus volume

With inhalation simulated at 15 L/min: for AR1, Δ*P* was -0.82 Pa; for AR2, Δ*P* was 2.33 Pa (see Fig. 1, Panels (g) and (h)). The hydraulic radius in AR1 was 3.43 mm, i.e. *ℛ* = 0.00343 m; the hydraulic radius in AR2 was 3.06 mm, i.e. *R* = 0.00306 m. Consequently using Equation 1, in AR1: the liquid bolus in each aspiration is *π* × *ℛ*^4^ ×|Δ*P*| */* (8 × 0.459 × 0.02) = 4.86 × 10^−9^ m^3^ = 0.00486 ml. Similarly in AR2: the liquid bolus volume ingested during each aspiration is 0.00874 ml.

### 3.2 Estimation of aspiration frequency and validation of the computational predictions

Using the earlier observational data in the first paragraph of Section 2.1, in conjunction with the computed volumes in Section 3.1, the aspiration frequency during each sleep cycle ranges between 0.021 ml*/*0.00874 ml and 0.021 ml*/*0.00486 ml, or approximately 2 – 4 times. Calculations based on direct experimental observations provide an estimate of 3 aspirations during a sleep cycle (derived in the third paragraph of Section 2.1, using previous reports^6,10^). The computational framework thus provides strong agreement with the direct experimental observations, providing a measure of validation for the underlying fluid dynamics framework.

### 3.3 Estimation of virion flow to the lower airway

Section 2.1 shows that the total number of aspirations of liquid boluses into the lower airway approximately ranges between 63 – 87 times in a day (including the subdued phase during sleep). Based on the data from the current subjects in the numerical simulations (see Section 3.1), the total volume of aspirated liquid in a day is thus between 0.00486 × 63 ml and 0.00874 × 87 ml, or between 0.3 – 0.76 ml. With the virological data from Section 2.3, the number of virions penetrating into the lower airway, while suspended in nasopharyngeal fluid boluses, is therefore between 0.3 × *𝒱*_ave_ = 2.1 × 10^6^ and 0.76 × *𝒱*_ave_ = 5.3 × 10^6^, while considering the mean viral load. For peak viral load (*𝒱*_peak_) in the sputum, the corresponding number of penetrating virions ranges over 7.1 × 10^8^ – 17.9 × 10^8^.

## 4 Discussion

We have demonstrated a novel computational approach to estimate virion flow rates to the lower airway from the initial dominant infection zone in the upper airway, i.e. the nasopharynx, which acts as the seeding site for the progressive infection via aspiration of virus-laden boluses of nasopharyngeal fluids. The computational fluid mechanics prediction of the aspirated liquid volume in each occurrence is validated by comparing the number of projected aspirations (approximately 2 – 4) during an eight-hour sleep cycle with prior observational findings of 3 aspirations in human subjects.

The numerically projected aspiration volumes have been linked to earlier records on aspiration frequency for the entire day, to obtain a total aspirated nasopharyngeal liquid volume of 0.3 – 0.76 ml each day. Subsequently using sputum assessment data from hospitalized COVID-19 patients, we see that the number of virions penetrating the lower airway ranges over 2.1 × 10^6^ – 5.3 × 10^6^, for mean sputum viral load. When the viral load peaks, the number of penetrating virions increases to 7.1 × 10^8^ – 1.8 × 10^9^. Deep sequencing studies suggest that the minimum infectious dose for SARS-CoV-2 transmission from one human host to another is on the order of *𝒪* 10^2^ virions^25^ (also supported by our earlier computational findings^8^), so it stands to reason that a dose 10^4^ – 10^7^ times higher will suffice to seed a second infection site within the same host, particularly given the relatively high levels of ACE2 expression in a subset of alveolar cells^26^.

Our results should be interpreted as being preliminary, given that the numerical findings are based on simulated data from only two CT-normal subjects. We note that there is good agreement between numerical predictions of aspiration frequency with earlier observational findings, which speaks to the validity of the underlying computational and mathematical framework. The size of the projected viral dose also speaks to the robustness of the conclusion – it is likely that virion flow from the nasopharynx to the lungs occurs in large excess of the minimum dose required to seed a lung infection for many individuals.

Our findings suggest a simple aspiration-based physiological mechanism for COVID-19 etiology following initial SARS-CoV-2 infection in the nasopharynx. Such a mechanistic link may be valuable in identifying risk factors that predispose patients to progress to COVID-19 disease following SARS-CoV-2 infection. For example, a prediction that flows readily (pun intended) from this proposed mechanism is that individuals with dysphagia (problems initiating swallowing) may be at increased risk of developing COVID-19 following SARS-CoV-2 infection, and may have more negative outcomes with the disease.

One condition associated with dysphagia is obstructive sleep apnea (OSA)^27–29^, with increased nocturnal aspiration^30^ and risk of aspiration pneumonia^31^. Based on the mechanism proposed by us for viral spread to the lungs, individuals with OSA would thus be expected to be at a higher risk for COVID-19. In fact, this has been reported by a number of different investigators^32–36^. One study, for example, reported an association of OSA with increased risk for hospitalization (OR 1.65; 95CI (1.36, 2.02)) and respiratory failure (OR 1.98; 95CI (1.65, 2.37)) due to COVID-19, after adjusting for diabetes, hypertension, and body mass index^37^. As a practical matter, our work suggests that individuals with OSA should not suspend the use of their CPAP devices upon testing positive for SARS-CoV-2, as has been suggested by some practitioners^38^.

As the prevalence of dysphagia also increases with other factors such as increased age^39^, cancer treatment^40^, and Parkinsonism^41^, the physiological mechanism proposed here may also account for some of the documented increased risk of adverse outcomes^42–45^ in these groups. Further study is required to understand the importance of dysphagia in general as a predictive factor for adverse outcomes with COVID-19.

Our findings also point to the critical importance of reducing SARS-CoV-2 viral load in the early stages of the infection. Therapeutics and prophylactics that fail to prevent the virus from gaining a foothold in the nasopharynx will allow the escalation of the disease in at least a subset of the population, which may already be more vulnerable to adverse outcomes from the disease. If vaccinal prophylaxis does not provide sterilizing immunity, then vaccinated individuals infected with SARS-CoV-2 in the nasopharynx may be subjected to a steady stream of virions testing their immune system with potential immune-evading mutations. Such a scenario is of particular concern for antibody prophylactics and vaccines that target the spike protein, work by us^46^ and others^47,48^ suggests that the spike protein has a relatively high mutational tolerance, and is expected to readily generate immune-evading mutations when antibody-based prophylaxis becomes widespread. In this scenario, some proportion of patients may experience breakthrough infections as a result of evolutionarily-mediated resistance, and seed transmission chains of antibody- or vaccine-resistant SARS-CoV-2 within the population. Biomedical interventions that rely on limiting the spread of the virus, that has established itself within the body, may underestimate the systemic threat that nasopharyngeal SARS-CoV-2 infections pose to individual patients as well as to public health.

## Data Availability

The project has generated simulated, quantitative, de-identified data on the flow physics in respiratory physiology. The data-sets (including Fluent .cas and .dat files) and the numeric protocols; along with MATLAB codes, Wolfram Mathematica notebooks, and Microsoft Excel spreadsheets used for data post-processing - are available on-request from the lead corresponding author (SB).

https://drive.google.com/drive/folders/1OnEo_fPtSzIqG5_VLKpsg3L5ixG108cI

## Data availability

We have generated simulated, quantitative, de-identified data on the flow physics in respiratory physiology. The data-sets (including Fluent .cas and .dat files) and the numeric protocols; along with MATLAB codes, Wolfram Mathematica notebooks, and Microsoft Excel spreadsheets used for data post-processing – are available on-request from the lead corresponding author (SB).

## Acknowledgments

The work is the result of an ongoing collaboration between SB (Assistant Professor at SDSU and Visiting Scholar at UNC Chapel Hill) and AC (Chief Executive Officer at Fractal Therapeutics). Computing facilities at both SDSU and UNC Chapel Hill have been used for this project. The authors also acknowledge Prof. Sunghwan Jung (at Cornell University), Prof. Leonardo Chamorro (at University of Illinois at Urbana-Champaign), and Prof. Diane Joseph-McCarthy (at Boston University) for several fruitful discussions.

